# A concise, machine learning-based questionnaire that screens for insomnia and apnoea in the general population

**DOI:** 10.1101/2020.05.15.20096404

**Authors:** Yizhou Yu, Samantha Jackson, Erla Björnsdóttir, Charles Oulton

## Abstract

Poor sleep is a major public health problem with implications for a wide range of critical health outcomes, including cardiovascular disease, obesity, mental health, and neurodegenerative disease.^1,2^ The most prevalent sleep disorders are insomnia and sleep apnoea. While questionnaires aimed at detecting and quantifying sleep problems have been used for years and proven to be reliable,^3-6^ they are often very extensive and scientifically worded. Here, we propose that the general population can use the SleepHubs Check-up (SHC), a concise questionnaire as a screening tool for sleep apnoea and insomnia. We validated the SHC against widely-used sleep questionnaires. These include the Insomnia Sleep Index (ISI)^5^ for detection of insomnia risk, as well as STOP-Bang^3^ and Multivariable Apnoea Prediction Index (MAPI)^7,8^ for the detection of sleep apnoea risk. We built a multivariate linear model to predict the ISI score based on the SHC questions and obtained an R^2^ of 0.60. For the detection of sleep apnoea, we constructed a convoluted neural network to predict the risk of apnoea from the SHC questions, and obtained an accuracy of 0.91. The SHC is therefore a reliable and accessible tool for the detection of latent sleep problems in the general public. Future work will aim at increasing the input data to improve the accuracy.

## INTRODUCTION

Poor sleep can negatively impact mood, cognitive performance and quality of life by increasing physiological arousal and sub-optimal cognition.^9^ It has been suggested that sustained sleep disturbance, as a result of obstructive sleep apnoea^10,11^ and Insomnia,^12,13^ results in cognitive deficits leading to neurodegeneration and mental health issues.

Recent reports on the prevalence of poor sleep in adults show that more than 33% of the population experience poor sleep.^14,15,16^ In countries including China, United States of America, Brazil and India, the prevalence of OSA exceeded more than 50% of the population, and a majority of sufferers remain untreated.^11,22^ and this number has been steadily increasing.^23^ A patient may suffer from chronic insomnia for several years, while treatments are increasingly effective at improving daytime and psychological functioning and underlying sleep patterns.^18,24^ If an individual suffers with both OSA and insomnia - also known as comorbid insomnia sleep apnoea (COMISA) - the combined effects are worse than the sum of the individual issues.^25^ Treatment of obstructive sleep apnoea (OSA) may leave individuals still feeling unrefreshed with a likely reduction in continuous positive airway pressure (CPAP) compliance.^26^ In order to improve an individuals’ outcome, treatment approaches should initially target COMISA individuals’ insomnia first to remove the barrier to CPAP treatment for OSA.^27^

Current assessment and diagnosis of sleep disorders involve a combination of measures; interviewing patients and their bed partner, physical examinations and in some instances polysomnography.^17^ The interviewing process usually takes the format of administering a number of recognised questionnaires; the Insomnia Severity Index (ISI) which has been shown to be a reliable and valid instrument to detect cases of insomnia in the population,^18^ the Epworth Sleepiness Scale which, in patients with obstructive sleep apnoea, significantly correlates with the respiratory disturbance index as recorded by overnight polysomnography.^4^ These practices are time consuming for both the individual and medical professionals and, combined with a lack of public awareness, can result in a number of undiagnosed sleep disorders. The increased number of people complaining of poor sleep puts a strain on doctors who claim that they lack sleep-related training.^19^

There are a number of easy, non-invasive, home sleep diagnostic tools and corresponding treatments available. In the case of obstructive sleep apnoea (OSA), pulse-oximetry and polygraphy can be used to diagnose, and CPAP^20^ or mandibular advancement device (MAD)^21^ are effective treatments. The availability and usage of consumer sleep technology (CST) and treatments for sleep disorders is increasing. While there are effective CST available to treat sleep problems, sleep questionnaires designed to characterise these problems are relatively complex, scientifically worded, and not easily accessible from the general public’s point of view.

This paper documents the validation of the SleepHubs Checkup (SHC). The SHC is a short, device-based questionnaire designed to engage and screen the general population to gauge if they have a sleep problem. It is quick and easy to complete and focuses on three categories commonly associated with poor sleep: daytime sleepiness, snoring, and awakenings including difficulties initiating and maintaining sleep. The questions used in the SHC took their inspiration from the Screen for Sleep Disorders.^28^ We chose questions for their brevity and simplicity and we adapted the wording and added a few supplementary questions to improve the accuracy of the screen. We designed several models which can accurately predict the ISI, MAPI and STOP-Bang scores.

## MATERIALS and METHODS

We randomly recruited 1198 healthy adults to answer questions on the STOP-Bang,^3^ ISI5 MAPI7 and the SHC (available at SleepHubs.com). Individuals readily participated, opting in at the start of the process, consenting to the use of anonymised data collected, by reading and accepting Sleep Hubs privacy policy. No incentive was given.

We recorded their responses to the four main questions 1. How do you feel during the day? 2. Are you happy with your sleep? 3. Do you ‘nod off’, doze or fall asleep during the day without obvious reason? 4. Has anyone ever complained about your sleep? and any resultant sub questions as part of the SHC.

The responses to STOP-Bang, ISI and MAPI questionnaires were recorded and scored according to their published validated scales and scoring mechanisms. Data analysis was performed by generating multivariate linear models and convolutional neural networks using TensorFlow (tensorflow.org) on the programming language R (R-project.org) to examine the consistency, reliability and validity of the SHC as an instrument to screen for potential sleep issues. We designed the neural network model to map the SHC to the validated questionnaires: MAPI and STOP-Bang (Snoring, Tiredness during daytime, Observed apnea, high blood Pressure, Body mass index, Age, Neck circumference, Gender) to distinguish whether an individual had a risk of insomnia and/or sleep apnoea. We trained the neural network on using a batch size of 128 and iterated the learning process for 2000 epoch. We then evaluated the neural network using 20% of the data, which the network has not previously seen. Finally, we trained a neural network using all of the data and uploaded that model onto our website SleepHubs.com for use by the general public.

## RESULTS

We first explored the gender and age split in our cohort (Figure 1). There were more than twice as many female participants as there were male ones. We noticed this gender disparity and sent an additional request to answer our questionnaire specifically targeted at male subscribers of SleepHubs. However, there were still more female respondents. We hypothesise that female patients may be more concerned by their sleep health.

**Figure 1.**
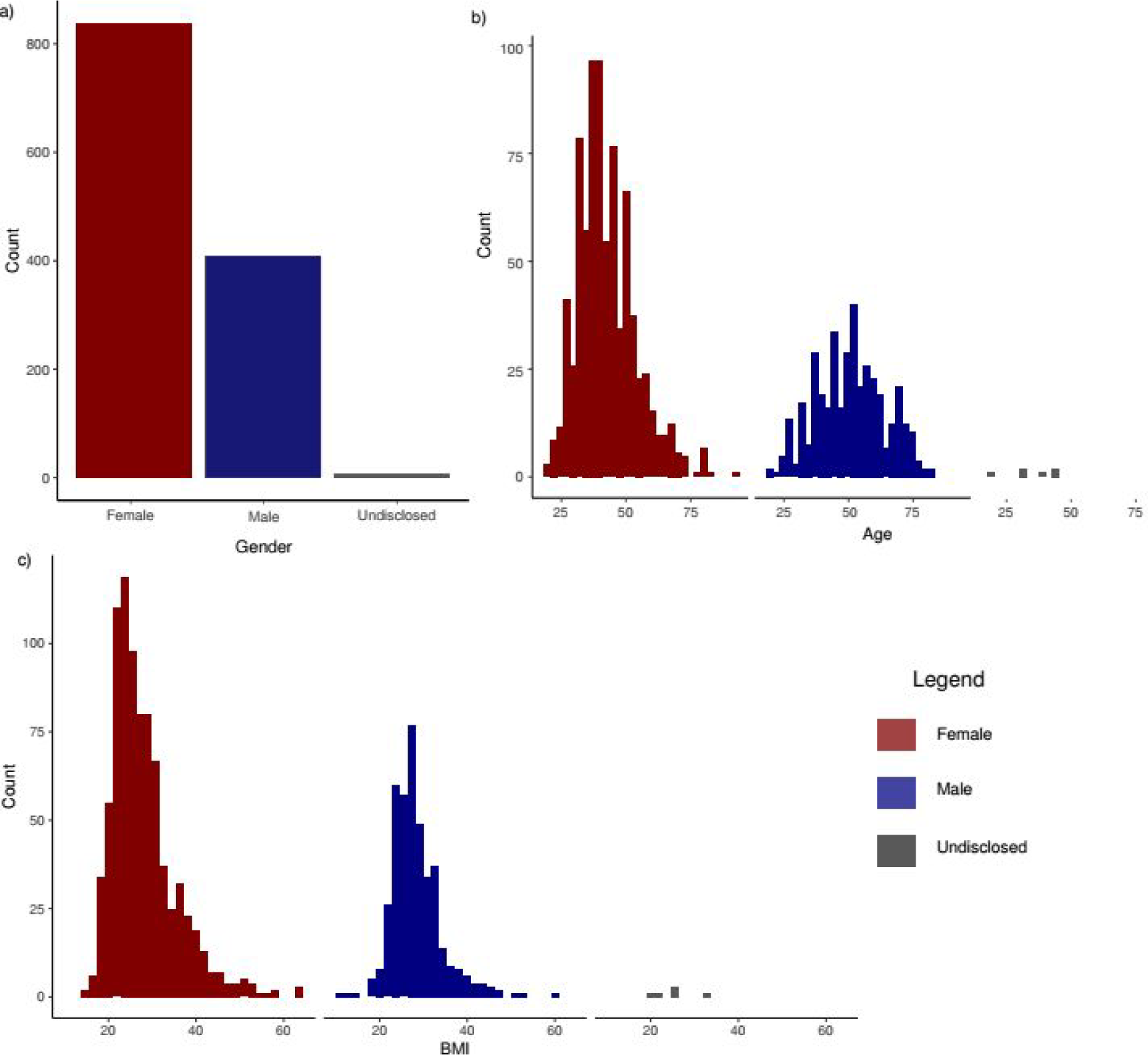
Descriptive statistics of the cohort. a. There are 838 females, 409 males and 6 individuals who do not wish to disclose their gender. b. Age distribution shown by gender, with the mean age value at 44.54±12.93 years. c. The body-mass index (BMI) distributions by genders, with the mean at 28.09±6.49.

Initially, a multivariate linear model was applied to the SHC questionnaire results in relation to the individuals corresponding ISI score (Figure 2a). The multivariate model of the SHC reliably predicts the probability of Insomnia with a relatively high accuracy (Figure 2a). This type of model, however, is unable to reliably predict the risk of OSA, with a low R^2^ approximately 0.30 (Figure 2b and 2c). The OSA models were nonetheless significant *(p-value* < 0.0001) and have a low residual standard error, prompting further investigation.

**Figure 2.**
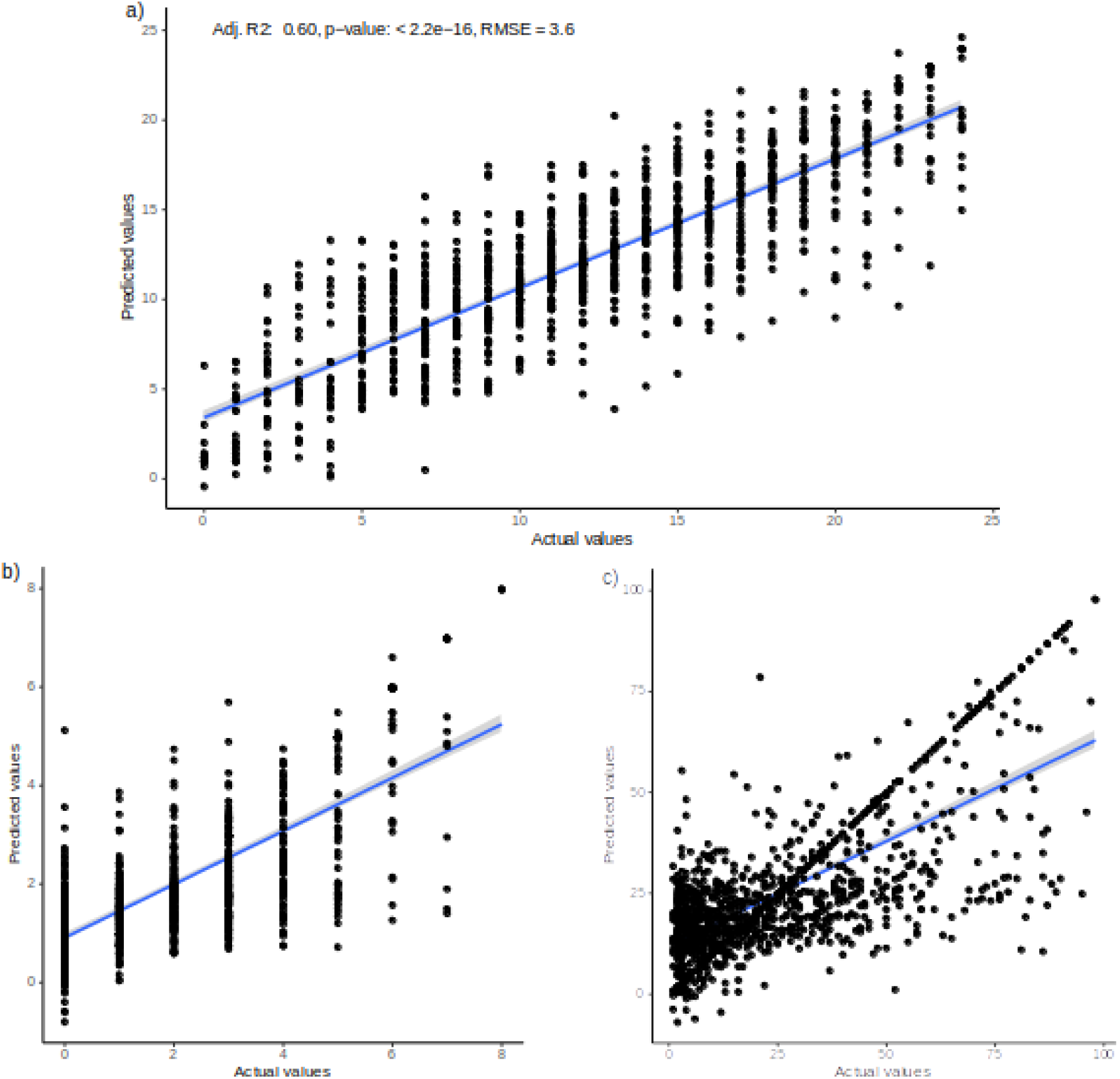
Multivariate linear models to predict the risk of insomnia using the ISI score (a) as well as OSA using StropBANG (b) and MAPI (c) The model that predicts ISI is a good fit, with an adjusted R^2^ of 0.60, *p-value <* 0.0001 and residual standard error of 3.6 on 827 degrees of freedom. The STOP-Bang and MAPI models are both significant (*p-value <* 0.0001), but have a low R^2^ of 0.34 and 0.31 respectively. The details and estimates of all 3 models are appended to this study as Supplementary Table 1.

We designed a convolutional neural network to predict the risk of OSA using both STOP-Bang and MAPI criteria, from the SHC questions (Figure 3). For mapping against STOP-Bang, our increased accuracy reached 0.76 (Figure 3a). In order to determine whether an individual is at the upper, middle or lower end of the STOP-Bang score, we rescored the data based on previous research criteria.^29^ The evaluation using the testing dataset yielded an optimal accuracy of 0.92 (Figure 3b).

**Figure 3.**
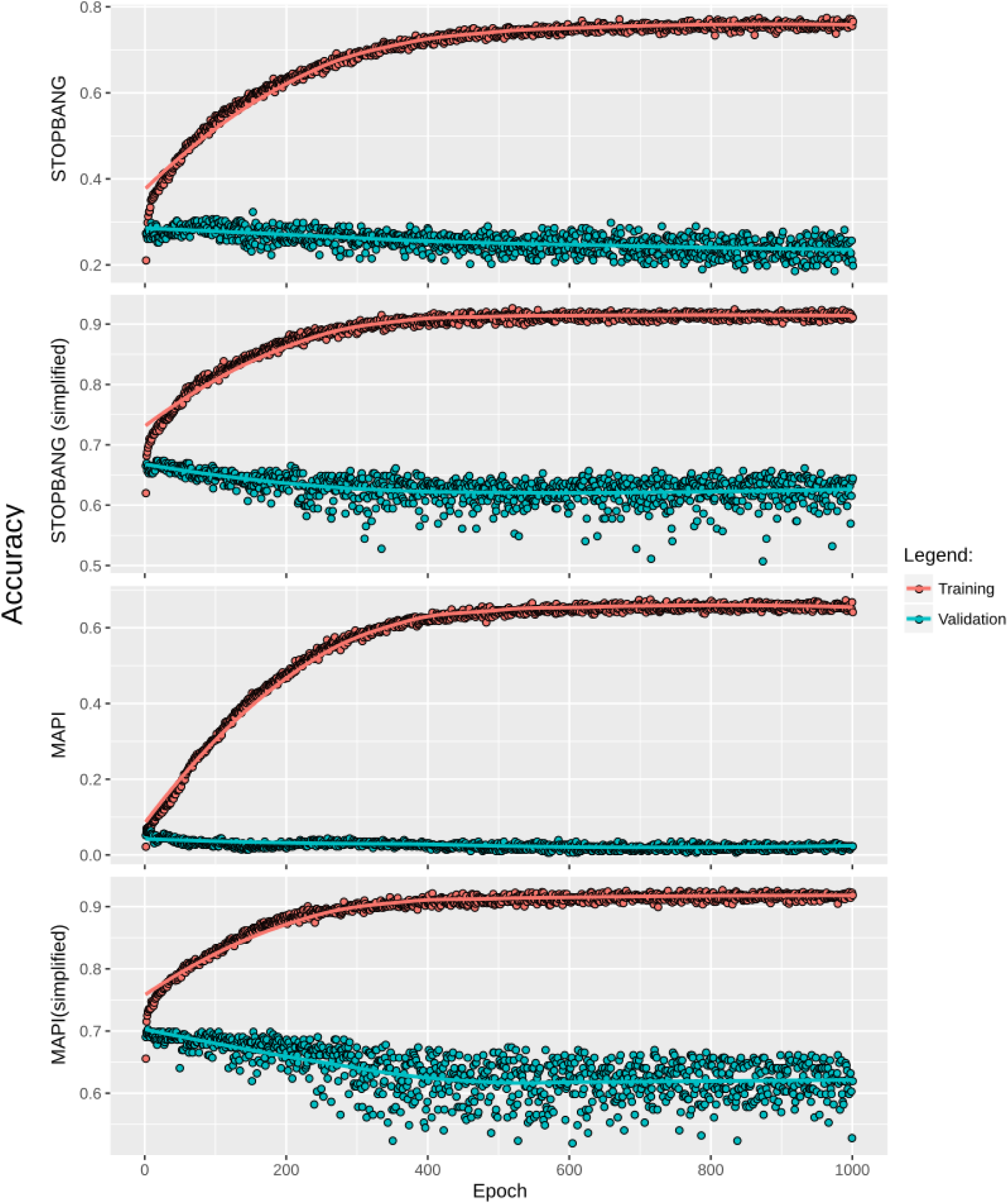
Neural network training results of the capability of the SHC of predicting OSA. The capability of predicting the exact STOP-Bang score from the SHC (a) and its approximate range under the groupings 0-2, 3-5, 6-8 (b). The capability of predicting the exact MAPI score from the SHC (c) and its approximate range under the groupings 0-33.3, 33.3-66.6, 66.6-100 (d). The accuracies of the predictions for both STOP-Bang and MAPI are increased to a maximum of 0.92 and 0.91 respectively.

A second neural network was trained to generate a MAPI score using the SHC answers. This score yielded an accuracy of 0.67 (Figure 3c). We then used a similar method to divide the MAPI scores into 3 groups of low, medium and high risk of apnoea scores and re-trained the neural network. The optimal accuracy reached was 0.91 (Figure 3d).

These results show that machine learning, using the convolutional neural network model, can reliably predict the risk of OSA from the SHC questions. The range of accuracies indicate that additional data will increase the accuracy of these models.

## DISCUSSION

Using data from 1198 healthy participants, we developed algorithms to predict the risk of sleep apnoea and insomnia based on 4 questions – the SHC. We calibrated the SHC against previously validated questionnaires, and showed that it can reliably distinguish whether an individual is at risk of insomnia and/or sleep apnoea. The strength of the SHC is that it is easily accessible for the general population.

In addition to its simplicity, the SHC can also be used by epidemiologists in the healthcare environment to detect the risk of the co-occurrence of both insomnia and OSA. COMISA is a relatively less investigated condition, and recent reviews call for better tools for its screening and diagnosis.^25^ The comorbid prevalence rates of between 30-50% of individuals that suffer from sleep disturbance have been highlighted throughout the last decade.^26,27,30^ Future research in these areas can use the SHC as an epidemiological tool to reliably screen for the comorbidity.

The main limitation of our study is that our cohort is predominantly female. This implies that our algorithms are currently more adapted to this population. It is therefore not recommended as an instrument to be used for clinical diagnosis, but it can be used easily in a time efficient manner as a valid instrument to screen for potential sleep issues.

In summary, the SHC is an inherently dynamic questionnaire as our machine learning algorithm will continue to improve with greater use. Researchers and clinicians are encouraged to use the questionnaire to further improve the algorithm. The SHC also has opportunities to map against other sleep disorders. Further research will elucidate the reliability of the SHC in detecting the risk of these less common sleep disturbances. Copyright SleepHubs Ltd. 2020 – This work is and remains the intellectual property of SleepHubs Ltd. (a company registered in England under GB10520947)

## Data Availability

Queries for data and code access can be directed to

## Ethics statement

The Medical Research and the Health Research Authority stated that our study does not require NHS REC approval for sites in England, Scotland, Wales and Northern Ireland.

## Acknowledgement

We thank Dr N. Stanley for helpful discussions.

**Abbreviations**
SHC: SleepHubs Check-up
ISI: insomnia severity index
MAPI: multivariable apnoea prediction index
OSA: obstructive sleep apnoea
MAD: mandibular advancement device
CPAP: continuous positive airway pressure
CST: consumer sleep technology

